# FAMILY-BASED RNA SEQUENCING IN BIPOLAR DISORDER FOR CANDIDATE GENE AND PATHWAY IDENTIFICATION

**DOI:** 10.1101/2025.01.30.25321384

**Authors:** Inés García-Ortiz, Miriam Martínez-Jiménez, Tomas Kavanagh, Lee L Marshall, José J Lucas, Peter R Schofield, Philip B Mitchell, Antony A Cooper, Janice M Fullerton, Claudio Toma

## Abstract

**Importance:** Genetic risk variants associated with bipolar disorder (BD) are likely to result in transcriptomic changes. RNA sequencing (RNAseq) studies in multiplex BD families are limited.

**Objective:** To identify candidate genes and pathways involved in BD.

**Design, settings, and participants:** This cohort study includes Australian families with three or more members diagnosed with BD. RNAseq was performed on total mRNA from lymphoblastoid cell lines of eight multiplex BD families. Patients were enrolled between May 1999 and October 2004. Data were analysed from June 2022 to March 2024.

**Exposures:** Differential gene expression changes and co-expressed gene networks in affected and unaffected relatives.

**Main outcomes and measures:** Quantified read counts were examined for differential gene expression and Weighted Gene Co-expression Network Analysis (WGCNA). Differentially Expressed Genes (DEGs) were validated through: i) gene-based association using BD Genome-Wide Association Study (GWAS) summary statistics; ii) Polygenic Priority Score (PoPS); and iii) replication using RNAseq data from brain tissues of 71 BD and 252 controls. DEGs and co-expressed modules were examined for enriched categories via Gene-Set Enrichment Analysis (GSEA) and Over-Representation Analysis (ORA), and were further validated through gene-set analysis using GWAS data.

**Results:** Sixty significant DEGs were found after comparing 16 BD (37.5% female) and 15 unaffected relatives (46.7% female), with the long non-coding RNA (lncRNA) *LINC01237* being the most significant. Gene expression patterns of DEGs were correlated between lymphoblastoid cell lines and brain tissues (r=0.83). After validation, five DEGs were prioritised, including *ENSG00000279277*, a lncRNA mapping in a GWAS locus for BD. Enrichment analyses of DEGs pointed to nervous system and gated channel activities. WGCNA identified five expression modules associated with BD. The most significant module showed enriched categories associated with ion transmembrane transport and hypoplasia on *corpus callosum,* together with a protein-protein interaction network related to solute carriers at plasma membrane.

**Conclusions and relevance:** This study implicates a role for lncRNAs in the pathophysiology of BD. Alterations in ion homeostasis, driven by the dysregulation of gated channels, appear to be a plausible impaired process in BD. In addition, the *corpus callosum* emerges as a key brain structure with a potential involvement in BD.

**KEY POINTS:** *Question:* Can transcriptomic signatures in families with multiple members affected with bipolar disorder (BD), likely due to a higher genetic load, identify genes and pathways contributing to disease risk?

*Findings:* In this cohort study of eight multiplex BD families, transcriptomic data implicated a pathogenic role of long non-coding RNAs, highlighting dysregulation of ion homeostasis as a key mechanism in the pathophysiologic aetiology of this condition.

*Meaning:* Non-coding RNA genes may play a more significant role in BD pathogenesis than previously estimated. The dysfunction of ion channels in BD indicates an endophenotype likely associated with channelopathy.

## INTRODUCTION

Bipolar disorder (BD) is a chronic psychiatric condition characterised by periods of mood fluctuation between manic and depressive phases.^1^ BD affects 1–2% of the adult population, and typically has an onset in early adulthood.^2,3^ Genetic factors play a substantial role in the aetiology of BD, with heritability estimates of approximately 80%.^4^ Genome-wide association studies (GWAS) are powerful approaches for the identification of susceptibility risk alleles. Recently, the Psychiatric Genomics Consortium (PGC) reported 64 genome-wide significant loci and additional suggestive loci, underlying that BD is a highly polygenic disorder.^5^ However, GWAS typically identifies common genetic variants of small effect sizes, which only explain between 20% to 30% of the genetic liability for BD.^5,6^ Whole Exome Sequencing (WES) and Whole Genome Sequencing (WGS) have provided insights into the genetic architecture of BD, indicating the additional contribution of rare genetic variants to the genetic liability of the disorder.^7^ Numerous studies have been recently performed to identify rare variants with higher penetrance effects segregating in families with multiple affected individuals.^8–11^ However, findings from family-based sequencing studies suggest modest penetrance of rare variants.^9^ Therefore, BD likely results from a complex interplay between hundreds of common risk alleles, several pathogenic rare variants, and *de novo* variants, each of which are observed both in sporadic cases and in families with multiple affected individuals.^7,9,12,13^ A limitation of GWAS meta-analyses and DNA sequencing studies is that each investigates only a portion of the complex genetic architecture of BD, often disregarding complex interactions amongst genetic risk variants, environment and epigenetic effects.

Transcriptomic studies through RNA sequencing (RNAseq) provide powerful approaches for capturing the effects of different classes of genetic variants and their interactions, providing the opportunity to identify potential convergence into pathophysiological processes relevant to disease. RNAseq studies have been performed in BD on brain tissues, whole blood or lymphoblastoid cell lines,^14–23^ and have employed either cohorts of patients with BD only,^14,20^ or comparisons between BD cases and controls.^16–19,23^ RNAseq studies in patients with BD cohorts only have investigated gene expression differences in relation to lithium responsiveness,^20,22^ or mood state,^14^ whereas case-control studies have explored differentially expressed genes (DEGs),^17–19,23^ or changes in gene co-expressed networks.^17,18,21^ Analysis of DEGs has suggested several candidate genes for BD, including those involved in ion channels,^17^ immune response genes,^18^ transcription factors, and G-protein-coupled receptors.^19,23^

Subtle gene expression changes may be missed when genes are inspected individually at the genome-level, but can be captured when acting synergistically within specific pathways under co-expressed modules. Indeed, gene co-expression network analyses have identified several candidate pathways potentially dysregulated in BD, such as mTOR signalling,^21^ immune response signal transduction,^17^ and regulation of cation channel activity.^17^

While numerous RNAseq studies have been conducted in BD to date, none have employed family-based approaches, which have the potential to enrich for genetic drivers of disease risk. An exception is a family-based study that compared iPSC-derived neurons from two BD-affected brothers with both their parents, implicating *WNT* signalling and ion channel subunits.^24^

In the present study, we perform RNAseq of immortalised lymphocytes in a cohort of eight multiplex families, including 16 affected and 15 unaffected relatives, aiming to identify novel candidate genes and pathways implicated in the disorder through differential gene expression analysis (DGE) and weighted gene co-expression network analysis (WGCNA), together with additional genomic analyses for prioritisation and validation.

## METHODS

### Study cohort and clinical assessment

This study was approved by the University of New South Wales Human Ethics Committee, and participants provided written informed consent. This study follows the Strengthening the Reporting of Genetic Association Studies (STREGA) reporting guideline. Eight nuclear families, each with at least three affected individuals across two generations, were selected from a larger cohort of 65 multiplex BD families. The selected families included individuals diagnosed with bipolar disorder type-I (BDI), bipolar disorder type-II (BDII), schizoaffective disorder-manic type (SZMA), and recurrent unipolar disorder (RUD) (Figure 1). All participants were clinically assessed between May 1999 and October 2004, via the administration of the “Family Interview for Genetic Studies” (FIGS) and the “Diagnostic Interview for Genetic Studies” (DIGS).^25,26^ Additional information is provided in eMethods in Supplement 1.

**Figure 1.**
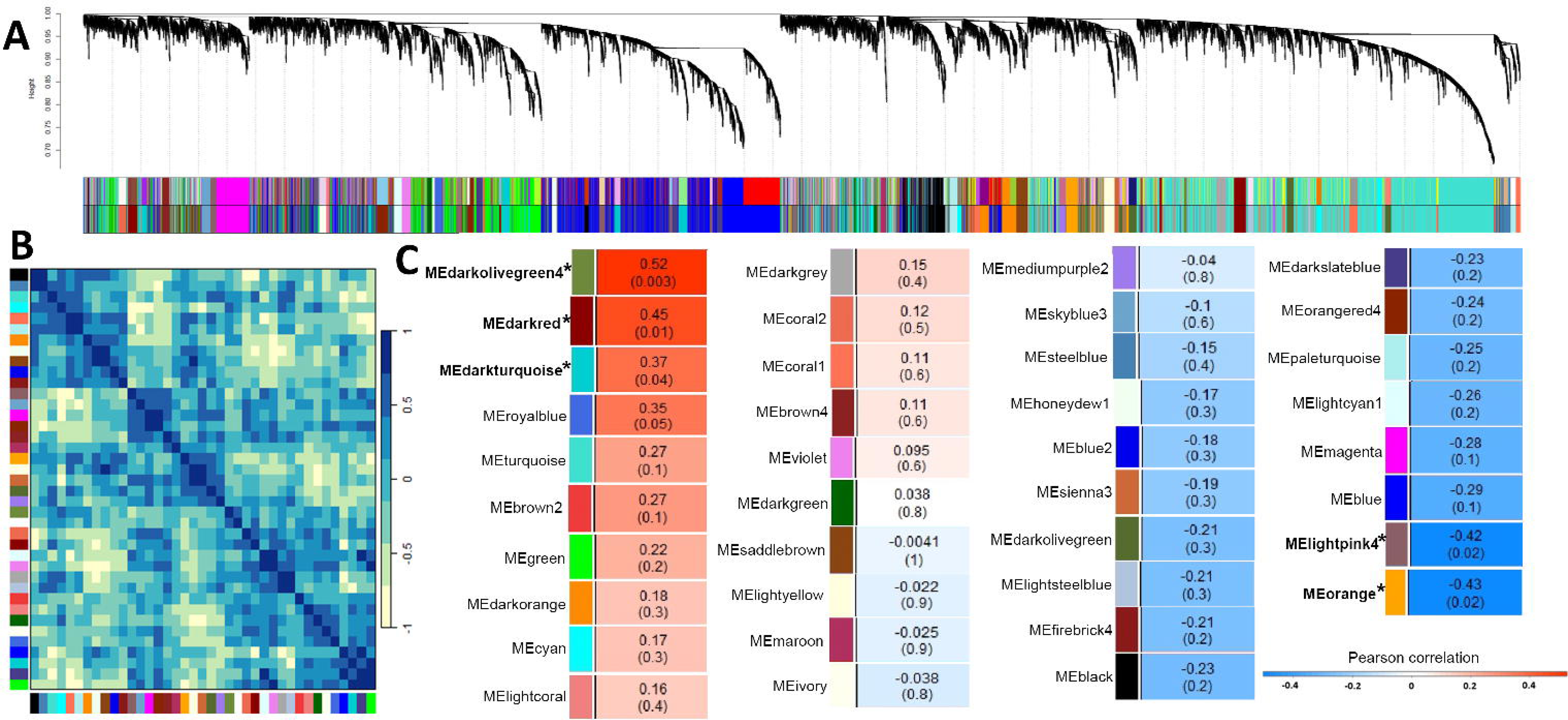
Structure of the eight multiplex BD pedigrees. Each family is depicted with multiple relatives whom underwent RNAseq (16 patients with BD and 15 unaffected relatives). Males are indicated with squares, females with circles, and psychiatric diagnoses are shown by dark shading as follows: i) full dark shading: bipolar disorder type I (BD-I); ii) full grey shading: bipolar disorder type II (BD-II); iii) bottom black shading: schizoaffective disorder-manic type (SZMA); iv) grey chess-board shading: recurrent unipolar depression (RUD); v) unshaded: phenotypically assessed unaffected relatives; v) question mark: diagnosis unknown. Individuals included in the RNAseq are marked with a star.

### RNA sequencing

Total mRNA was extracted from lymphoblastoid cell lines of 31 individuals and rRNA depleted using RNAseH. Libraries were prepared with Illumina Tru-seq stranded mRNA library kit (following manufacturer’s protocol). Prepared libraries were amplified for 13 cycles using the KAPA HiFi HotStart Library Amplification Kit (Roche). Libraries were then sequenced on the Hiseq 2500 (Illumina) at The Kinghorn Centre for Clinical Genomics (Garvan Institute of Medical Research, Sydney, Australia) using paired-end sequencing v4.0 chemistry over 125 cycles.

### RNAseq Data Preprocessing

Raw reads were quality checked using FastQC version 0.11.9 (http://www.bioinformatics.babraham.ac.uk/projects/fastqc), trimmed using Trimmomatic version 0.39,^27^ and then rechecked for quality. Alignment was performed using Hisat2 version 2.1.0 against the GRCh38.p13 reference genome.^28^ The resulting mapped bams were then sorted and indexed using samtools version 1.9.^29^ Gene quantification was performed using HTSeq version 2.2.1.^30^ Additional information on principal component analysis (PCA) and MA plot is provided in eMethods. The complete workflow is shown in eFigure 1 in Supplement 1.

### Gene Expression Analyses

The workflow and additional methodological details of the present study are provided in eFigure 2 and eMethods in Supplement 1, respectively. The DESeq2 package (version 1.36) was used in R (version 4.3) to perform DGE analysis between gene read counts of patients with BD and their unaffected relatives.^31^ The DGE analysis model included ‘sex’, ‘age’ and ‘family ID’ as covariates.

**Figure 2.**
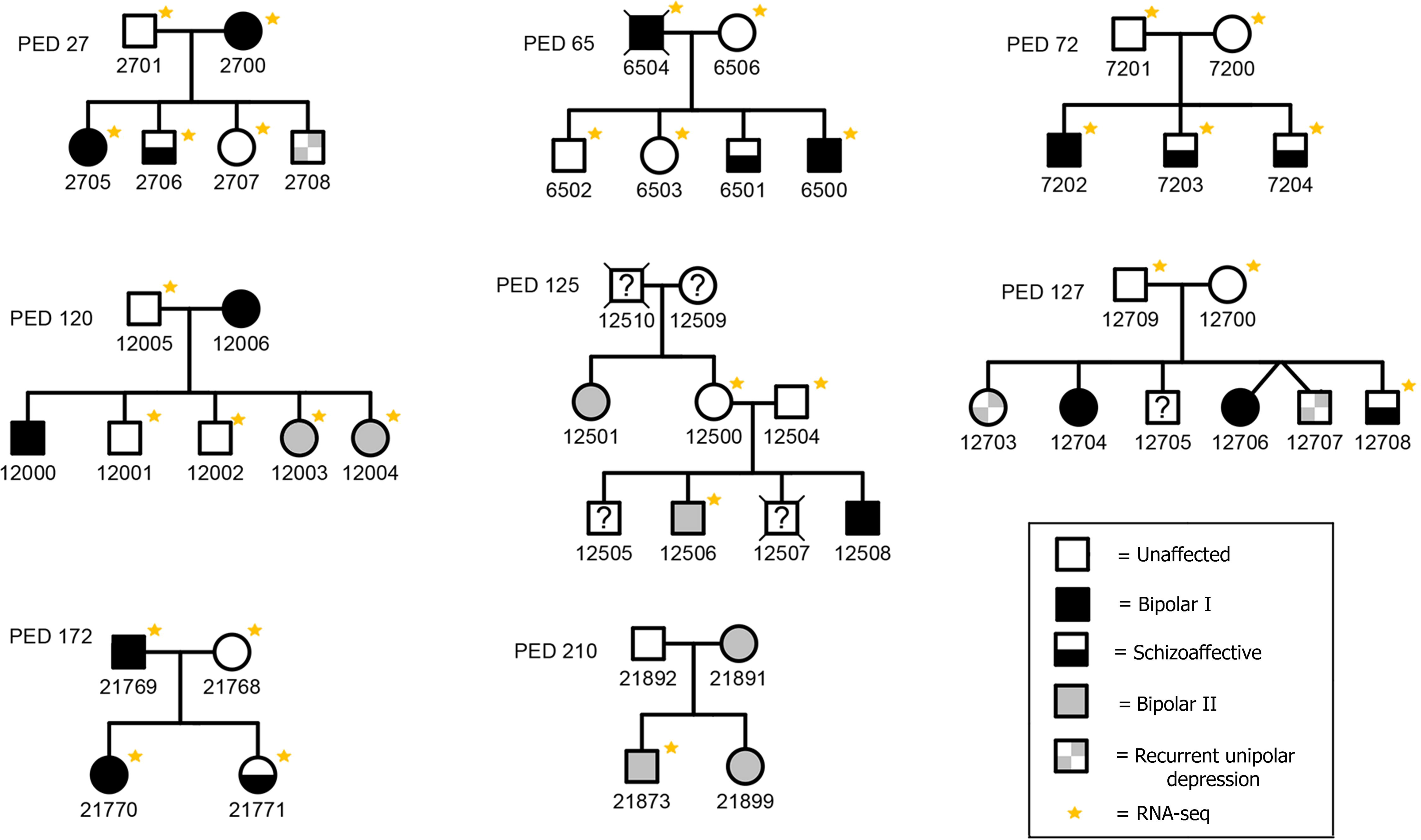
DEGs and validated enriched categories. A). Volcano plot shows differential gene expression analysis of 15,231 genes. The 60 DEGs (adjP<0.05) are depicted above the dashed line (threshold for significance: *P* <1.9E-04; -log10 *P* > 3.71). The DEGs with |log2 fold-change| > 0.3 are depicted in red and labelled with their gene symbol. B). GSEA categories validated by MAGMA GSA. Each category is represented by the enrichment Z-score (X-axis) and P-value (Y-axis). Bubble area represents the set size, and colour represents the database source.

Raw data from the Genotype-Tissue Expression (GTEx) database version 8 (https://gtexportal.org) were used to assess gene expression correlation between 13 brain tissues, whole blood and EBV-transformed lymphocytes for the differentially expressed genes (DEGs) of our study.

A signed gene co-expression network was constructed using the weighted gene co-expression network analysis (WGCNA) version 17.1 package in R.^32^ The count matrix was filtered to discard gene features with fewer than ten reads in 90% of the samples. The adjacency matrix was built by applying the signed similarity matrix to a soft threshold power (β), chosen according to the scale-free topology criterion and set to 12 (r^2^ cutoff = 0.8). The correlation between the resulting modules and affection status was assessed using the Pearson correlation test. The module most strongly correlated with the trait was further investigated for gene network relationships and in depth interaction analysis using the Ingenuity Pathway Analysis (IPA) software release 2023 (www.ingenuity.com).

### Enrichment Analyses and Validation

The DGE results were examined for enriched categories using the Gene-Set Enrichment Analysis (GSEA) through the *fgsea* function in R,^33^ while associated modules from WGCNA were inspected for enriched categories via Over-Representation Analysis (ORA) through the *enricher* function in R.^33^ The enrichment analyses from DGE and WGCNA were performed using MSigDB in R using the following databases: Gene Ontology (molecular function, biological processes, cellular component), Human Phenotype Ontology (HPO), Reactome and the Kyoto Encyclopaedia of Genes and Genomes (KEGG).^34^ Adjusted *P-values* were calculated for each enriched category, with a significance threshold set at 0.05.

The DGE results were validated as follows (eFigure 2 in Supplement 1): i) A replication study using an independent RNAseq dataset from the PsychEncode Consortium BrainGVEX RNAseq Study, which after quality control, included 71 patients with BD and 252 controls with RNA derived from human brain tissue (BA9);^35^ ii) Gene-based association analysis via MAGMA version 1.9,^36^ using PGC3 GWAS summary statistics of BD (41,917 BD; 371,549 controls);^5^ iii) Polygenic priority score (PoPS) version 2.0,^37^ which prioritizes candidate genes by combining genetic associations and functional annotations. Additionally, enriched categories identified from either GSEA or significant modules of WGCNA were validated through MAGMA gene-set analysis (GSA).

## RESULTS

### DGE and enriched categories

The raw count matrix initially consisted of 61,806 genetic features. After applying a first filter to include only genes with more than ten total reads across all individuals, the dataset comprised 28,525 features in total. PCA showed no clear clustering of subjects or obvious outliers (eFigure 3A in Supplement 1). Thus, all samples were retained for analysis. Following additional filtering of normalised read counts to reduce false-positive results (eMethods in Supplement 1), the DGE analysis was performed on raw read counts from a total of 15,231 genes. The MA plot showed the expected trumpet-shape distribution of expressed genes based on the log fold-change between the two groups and the means of normalised counts (eFigure 4 in Supplement 1). DGE analysis identified 60 DEGs (*adjP*<0.05) out of 15,231 genetic features (Figure 2A and eTable 1 in Supplement 2), with *LINC01237* being the most significant gene (*adjP*=4.1E-07). PCA using only the 60 DEGs effectively clustered the patients with BD from the unaffected relatives (eFigure 3B in Supplement 1). In a hierarchical cluster analysis, the gene expression patterns of the 15 most significant DEGs (*adjP*<0.01) successfully grouped the individuals based on their diagnostic status (eFigure 5 in Supplement 1). Enrichment for biological categories was performed using the Wald statistic as a ranking metric in the GSEA analysis for the 15,231 genetic features, resulting in 500 enriched categories (*adjP*<0.05; eTable 2 in Supplement 2). These categories underwent validation through a GSA using the available PGC3 BD GWAS summary statistics.^5^ This analysis identified 68 validated categories based on BD genetic associations (Figure 2B). The top significant categories included “neuronal system” (*P*=2.1E-06), “voltage gated cation channel activity” (*P*=3.1E-05), and “gated channel activity” (*P*=5.1E-04) (eTable 3 in Supplement 2).

### Candidate genes from the DGE study

To assess whether gene expression differences detected in lymphoblastoid cell lines are relevant to gene expression in the brain, we employed correlation analysis of the 60 significant DEGs across these tissues, using transcripts per million (TPM) data downloaded from the GTEx portal (eTable 4 in Supplement 2). The Pearson correlation coefficient (r=0.832) was calculated between the average TPM of 13 brain tissues and the lymphoblastoid cell line, showing a relatively high correlation. Out of the 60 DEGs, 55 showed TPM values greater than 0.1 across all brain tissues, indicating basal gene expression (eFigure 6 in Supplement 1). The validation of the 60 DEGs was performed using complementary approaches: i) a DGE analysis from an independent RNAseq dataset from brain tissues of 71 patients with BD and 252 controls (eTable 5 in Supplement 2); ii) MAGMA gene-based analysis, which identified 15 significant genes; and iii) PoPS analysis, which identified seven genes with scores greater than 0.2 (eTable 6 in Supplement 2). Table 1 lists high priority DEGs that achieved a positive validation in at least two of the three validation approaches. In this prioritization, we include also *ENSG00000279277*, a long non-coding RNA (lncRNA), which lacks a PoPS score as this metric is only applicable to protein-coding genes. This lncRNA maps to a genome-wide significant locus for BD (eFigure 7 in Supplement 1).

**Table 1.** List of high priority DEGs that fulfil at least two of the three validation criteria: *adjP*<0.05 in the replication dataset of PsychEncode, *P*<0.05 in the MAGMA gene-based analysis, PoPS score >0.2. This prioritization includes also the lncRNA *ENSG00000279277*, mapping on a GWAS significant locus for BD.

| **Ensembl Gene ID** | **Gene Symbol** | **PsychEncode (P)** | **PsychEncode (*adjP*)** | **MAGMA (P)** | **PoPS score** |
| --- | --- | --- | --- | --- | --- |
| ENSG00000229127 | *KANSL1L-AS1* | 4.3 E-04 | **0.0278** | **0.0027** | NA |
| ENSG00000135870 | *RC3H1* | 6.64 E-03 | 0.1038 | **0.0017** | **0.3376** |
| ENSG00000125534 | *PPDPF* | 0.0211 | 0.1812 | **0.0258** | **0.3140** |
| ENSG00000279277 | - | 0.8067 | 0.9244 | **1.59E-07** | NA |
| ENSG00000172354 | *GNB2* | 0.9260 | 0.9729 | **0.0051** | **0.790** |

NA, PoPS score is not computable for non-protein coding genes due to incorporation of protein-protein interaction data.

### WGCNA and correlated modules to BD

The initial 61,806 genetic features were filtered to discard genes with fewer than 10 read counts in 90% of the samples, which left 14,938 genes for the analysis. From the gene network, 38 modules were assembled, and five of them showed significant correlation with the trait (P<0.05) (Figure 3), with the *darkolivegreen* being the most significant (*P*=0.003) (eTable 7 in Supplement 2). A complete list of genes for each module, along with their module membership value is provided in eTable 8 in Supplement 2. ORA was performed considering the gene set from each of the significant module, resulting in 197 enriched categories (P<0.05; eTable 9 in Supplement 2). After MAGMA GSA, 13 categories were validated (Table 2), with the most significant being related to ion transmembrane transport categories and hypoplasia of the *corpus callosum*, all belonging to the *darkolivegreen4* module. The 98 genes of this module were inspected using IPA to identify relevant protein-protein direct interaction networks for BD. Interestingly, the top network identified several plasma membrane solute carrier (SLC) members, as well as genes associated either with BD or schizophrenia (eFigure 8 in Supplement 1).

**Figure 3.**
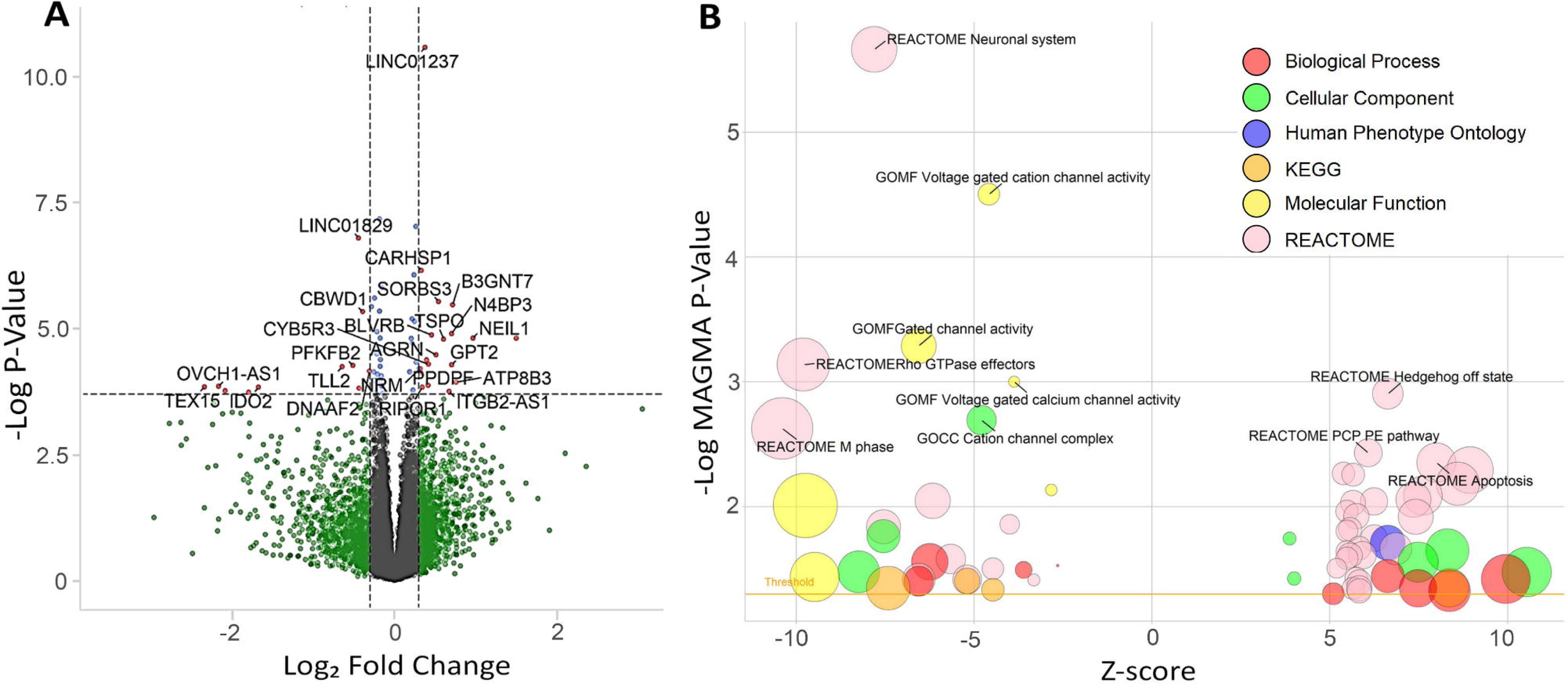
Weighted gene co-expression network analysis. A). Gene dendrogram (above) and module assignment (below) by Dynamic Tree Cut, which resulted in 38 assembled modules after merging modules with 75% of correlation. B). Heatmap of module eigengene correlations. C). Correlation of each eigengene module to BD. For each module (box) is shown the Pearson correlation coefficient and *P-value* between parentheses; positive correlations are depicted in red shades and negative correlations are depicted in blue shades, with stronger correlations in darker colours. The modules that are significantly correlated with BD are indicated with an asterisk and the module name is in bold text.

**Table 2.** Enriched categories via ORA are shown per module after validation via gene-set analysis (GSA) with MAGMA.

| **ID Category (Database)** | **Module** | **GSA MAGMA** |
| --- | --- | --- |
| Cation transmembrane transporter activity (GOMF) | Darkolivegreen4 | 0.0012 |
| Cation transmembrane transport (GOBP) | Darkolivegreen4 | 0.0022 |
| Hypoplasia of the corpus callosum (HPO) | Darkolivegreen4 | 0.0051 |
| Ion transmembrane transporter activity (GOMF) | Darkolivegreen4 | 0.0098 |
| Condensed chromosome centromeric region (GOCC) | Darkturquoise,<br>Darkred | 0.0168 |
| DNA packaging complex (GOCC) | Darkturquoise | 0.0172 |
| Protein DNA complex assembly (GOBP) | Darkturquoise | 0.0180 |
| DNA replication initiation (GOBP) | Darkturquoise | 0.0301 |
| Protein DNA complex (GOCC) | Darkturquoise | 0.0333 |
| Protein heterodimerization activity (GOMF) | Darkturquoise | 0.0365 |
| Mitotic DNA replication (GOBP) | Darkturquoise | 0.0371 |
| Import across plasma membrane (GOBP) | Darkolivegreen4 | 0.0395 |
| Protein DNA complex subunit organization (GOBP) | Darkturquoise | 0.0399 |

Abbreviations: GOMF, GO Molecular Function; GOBP, GO Biological Process; GOCC, GO
cellular Component; HPO, Human Phenotype Ontology

## DISCUSSION

Patients with BD with a strong family history are hypothesised to have a higher genetic load than sporadic cases, and therefore have been posited as being useful for genomic studies. Families with multiple relatives diagnosed with BD have traditionally been employed for linkage studies, which have not provided the required resolution for identifying causal genes. More recently, several studies have performed WES or WGS in nuclear or extended families, which have started to unveil putative genetic mechanisms and susceptibility genes.^8–12^ Conversely, transcriptomic studies in multiplex BD families have been limited to one single family.^24^ In this study, we conducted a family-based RNAseq study using a cohort of eight BD multiplex families performing DGE followed by WGCNA, enrichment analyses and further validation via an independent case-control cohort, genetic associations and gene prioritization methods.

DGE analysis suggested a role in BD pathogenesis for lncRNAs. These master modulators are implicated in the regulation of a large numbers of biological processes, highlighting their potential significance in psychiatry.^38^ Although still largely unexplored, lncRNAs appear to be highly species-specific. Indeed, 8,000 more lncRNAs were identified in humans than mice, with half of them found only in the brain, suggesting that this class of genes may have played a role in human evolution, leading to a greater complexity and brain connectivity than other mammals.^39^ In our study, the most significant DEG is *LINC01237*, which has also been identified as a top differentially methylated region (DMR) between patients with BD and controls.^40^ Another report found a DMR in this gene modulating the response to lithium in BD.^41^ Following validation approaches, two additional lncRNAs, namely *KANSL1L-AS1* and *ENSG00000279277*, emerged as putative candidates for BD in our study. Notably, a genome-wide significant association at rs696366 was found within the first intron of *ENSG00000279277* in the meta-analysis GWAS of BD,^5^ suggesting the potential involvement of this lncRNA or the closest *CD47*, or both. Furthermore, our study pointed also to *RC3H1*, *PPDPF* and *GNB2* as potential candidate genes for BD. *GNB2* is involved in G protein-coupled receptor (GPCR) signalling transduction via GTPase activity. Recent transcriptomic studies in brain tissues have reported GPCRs as a protein category with a primary role in psychiatric diseases, particularly in BD.^23,42^ Additionally, *de novo* variants in *GNB2* are known to cause a syndromic form of intellectual disability.^43^ *PPDPF* is ubiquitously expressed, and participates in the mTOR signalling,^44^ which was found to be upregulated in a transcriptomic study of brain tissues in BD,^21^ although no previous genetic association has been reported. *RC3H1* binds to the 3’ UTR of mRNAs, leading to mRNA deadenylation and degradation, and plays a significant role in immune and inflammation processes.^45^ Alterations in neuroinflammation have been widely described in subgroups of psychotic patients.^17,18,46,47^

After validation, enrichment analyses of DEGs pointed to top significant categories related to gated channel activities and ion transport functions, particularly those associated with cation channels. These classes of proteins are essential for neuron excitability, including the action potential generation and signal propagation. ^17,48^ Dysregulation of cellular calcium homeostasis can alter neuronal excitability, potentially explaining mood episode fluctuations observed in BD.^48,49^ Numerous genetic studies in BD identified common risk alleles and predicted pathogenic rare variants in genes encoding neuronal ion channels, including voltage-gated calcium channels and sodium channel related genes.^11,50–52^ These findings collectively support the hypothesis of a molecular mechanism related to a channelopathy, at least in a subset of patients with BD, characterized by dysfunction of ion channels or their associated proteins.^49,53,54^

Other enriched categories, though less prominent, involve intriguing signalling pathways such as Rho GTPases, Hedgehog, and planar cell polarity represented by Wnt pathway. These pathways provide compelling candidates for biological mechanisms potentially implicated in the pathophysiology of BD.

While DGE accounts for changes in expression of individual genes, the WGCNA captures altered expression patterns of gene clusters that otherwise would be missed in the DGE analysis. WGCNA is particularly effective in a complex polygenic disorder such as BD, where the result of thousands of common risk variants of small size effects on gene expression may act in concert to compromise specific pathways. We identified five modules associated with BD, with the *darkolivegreen4* module showing the strongest association. Enrichment analysis of this module after validation pointed to ion transport across membranes, directly aligning with the findings from our DGE analysis on gated ion channels. Protein network analysis of this module identified several solute carrier proteins at plasma membrane and downstream interaction with associated genes with BD and schizophrenia.

Another significant enriched category corresponds to ‘hypoplasia of the *corpus callosum*’, a major nerve tract connecting the cerebral hemispheres. Interestingly, one of the largest neuroimaging studies, performed by the ENIGMA BD working group and involving over 3,300 individuals, identified the *corpus callosum* as key structure for the disorder, reporting lower fractional anisotropy and the highest effect sizes in patients with BD.^55^ Structural and functional abnormalities in the *corpus callosum* have been consistently reported in BD,^56–60^ suggesting that impaired interhemispheric connectivity, potentially caused by altered myelination, may play a critical role in the pathophysiology of the disease. Our findings provide further evidence of the putative role of the *corpus callosum* in BD.

## Limitations

The RNAseq data were derived from immortalized lymphocytes cell lines, which may not fully recapitulate the gene expression changes in brain tissues. However, for the significant DEGs identified in our study we found strong correlation between transformed lymphocyte cell line and brain tissue expression. Although our multiplex BD family cohort is relatively small, it represents the largest cohort used to date for RNAseq studies in this context. The PoPS validation approach is limited to coding genes, and does not currently account for genes on the sexual chromosomes.

## Conclusions

In this family-based transcriptomic study of multiplex BD families, we suggest that lncRNAs may play a primary role in the pathophysiology of BD. Through validation approaches of our significant DEGs, we identified several putative candidate genes, including *ENSG00000279277*, a lncRNA mapping to a GWAS significant locus for BD. Enrichment analyses of DEGs and WGCNA significant modules consistently pinpointed to abnormal patterns in gated channels and ion transmembrane transport. Additionally, enrichment analysis identified gene networks implicated in the *corpus callosum*, a brain structure recently emerging as prominently altered in BD.

## Authors Contributions

Dr Toma had full access to all of the data in the study and takes responsibility for the integrity of the data and the accuracy of the data analyses.

*Concept and design:* García-Ortiz, Martínez-Jiménez, Cooper, Fullerton, Toma

*Acquisition, analysis, or interpretation of data:* All authors

*Drafting of the manuscript:* García-Ortiz, Martínez-Jiménez, Toma

*Critical revision of the manuscript for important intellectual content*: All authors

*Statistical analyses:* García-Ortiz, Martínez-Jiménez, Fullerton, Toma

*Obtained funding:* Cooper, Fullerton, Schofield, Toma, Mitchell

*Administrative, technical, or material support:* Kavanagh, Marshall, Lucas-Lozano, Schofield, Mitchell, Cooper, Fullerton.

*Supervision:* Toma, Fullerton, Cooper

## Meeting Presentation

This study was presented, in part, at the Annual Meeting of the World Congress of Psychiatric Genetics; 9-14 October 2023, Montreal, Canada.

## Data Sharing Statement

See Supplement 3

## Additional Contributions

We thank Anna Heath at Neuroscience Research Australia (NeuRA) for transformed lymphoblastoid cell line establishment and handling.

## Data Availability

Individual-level data are not available for data sharing. Sharing of summary data (raw count matrix) will be considered upon request.

