## Supplement 1 for "FAMILY-BASED RNA SEQUENCING IN BIPOLAR DISORDER FOR CANDIDATE GENE AND PATHWAY IDENTIFICATION"

1. **eMethods**

1.6 DEGs validation…………………………………………………………………………...5

1.7 WGCNA pipeline………………………………………………………………………….7

2. **eFigures**

2.1 eFigure 1: Workflow of the RNAseq data preprocessing…………………………………9

2.2 eFigure 2: Computational workflow of the present study…………………………………10

2.3 eFigure 3: PCA plots of count matrix of genetic features in the total of 31 samples……..11

2.4 eFigure 4: MA plot………………………………………………………………………..12

2.5 eFigure 5: Heatmap of DEGs……………………………………………………………..13

2.6 eFigure 6: Gene expression patterns of DEGs across tissues……………………………..14

2.7 eFigure 7: Association plot of *ENSG00000279277* from BD GWAS……………………15

2.8 eFigure 8: Protein-protein interaction network of the *darkolivegreen4* module………….16

3. **eTables in Supplemental Online Tables (Supplement 2)**

eTable 1: The DGE analysis results comparing affected and unaffected individuals in eight multiplex bipolar families

eTable 2: Enriched categories from the gene-set enrichment analysis (GSEA) of the DEGs

eTable 3: Validation of the categories from the GSEA results via GSA

eTable 4: Comparisons of TPM expression data of the 60 DEGs across tissues of interest

eTable 5: DGE analysis from brain tissues of an independent cohort of 71 BD and 252 controls

eTable 6: The *in silico* validation of sixty DEGs using complementary approaches

eTable 7: Modules from the WGCNA correlated with BD

eTable 8: List of genes and module membership values from the WGCNA

eTable 9: Results from the ORA for each associated module and category validation

**1.1 Clinical assessments**

Medium to large multiplex and extended multigenerational pedigrees were collected through the Mood Disorders Unit and Black Dog Institute at the Prince of Wales Hospital, and the School of Psychiatry, University of New South Wales, Sydney, Australia. All families were ascertained after initial consultation with a proband with bipolar disorder type I (BD-I). Each pedigree member provided peripheral blood samples for DNA extraction and creation of cell lines by standard laboratory methods. Information obtained from the FIGS, DIGS, and medical records was used to generate best-estimate Research Diagnostic Criteria (RDC) for DSM-IV bipolar disorder type I (BD-I), bipolar disorder type II (BD-II), schizoaffective disorder-manic type (SZMA) or recurrent unipolar depression (RUD).^1^ All experiments were carried out following regulations and guidelines approved by the University of New South Wales Human Research Ethics Committee (initial approval HREC04144; extensions HREC10078, HC15503, HC16347).

**1.2 Cell culture and RNA extraction**

Cell lines were created using Epstein–Barr virus (EBV) transformation immediately after blood collection, and vials containing EBV-transformed lymphocytes were archived in liquid nitrogen. At the beginning of this experiment, vials were thawed in ~6 batches of 4-6 lines per batch; washed in 10ml PBS,centrifuged at 300G for 5min, resuspended in 10ml media (Dulbecco's Modified Eagle Medium media with 10% Fetal Calf Serum and 4mM l-glutamine; Gibco, Thermofisher), seeded into T25 flask in upright position and placed in a 37^o^C incubator with 5% CO_2_ [Passage 1; P1]. At P1-48 hours, media colour was checked, more media added if required and flasks were laid down. At P1-72 hours, cells and media were transferred to a falcon tube, centrifuged at 300G for 5min, resuspend and seeded at 2×10^6^ cells into 10ml media in T75 with flask lying down [Passage 2; P2]. At P2-96 hours [or 168 hours after thawing], growth rates and doubling time were calculated, and estimated at 41.74±8.45 hours (30.8-69.6 hours). At P2-96 hours [P2], cells were counted and reseeded at ~4×10^6^ cells into 10ml media in T75 [Passage 3; P3]. At every 24 hours after P2-96 hours, media colour was checked, cell clumps resuspended and fresh media added if required (n=5). At P3-96 hours [or 264 hours/11 days after thawing], approximately ~9×10^6^ cells were harvested into trizol. cDNA was synthesized from 3 μg total RNA per case using SuperScript^®^ III First-Strand Synthesis kit and random hexamers (Invitrogen).

**1.3 RNAseq Data Preprocessing**

A complete overview of read processing, alignment and gene read counts is shown in eFigure 1. The trimming process parameters were set to include reads with a minimum length of 50 bp and with the last 4 bp showing 20 or above in quality score. For quality scores lower than 20 in the last 4 bp, the last base pair was removed and then reassessed for these parameters. The trimmed reads were then re-checked in a second quality control with FastQC. The read counts quantification was performed using the intersection-strict mode in HTseq, which discards reads that do not fully overlap an exon.

**1.4 Differential Gene Expression Analysis**

The DESeq2 software used as input file the raw read count matrix that comprised 61,806 genomic features from both coding and non-coding genes from the Ensembl GRCh38.p13 reference genome. A first filtering step was applied to delete the features <10 count reads across the 31 samples. The model for this analysis was built taking into account ‘gender’, ‘age’ and ‘familyID’ as covariates. DESeq2 uses a negative binomial generalised linear model (GLM) represented as follows:

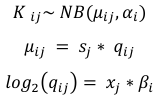

The counts (*K_ij_*) for gene *i* and sample *j* are adjusted into a negative binomial distribution with mean *𝜇_ij_* and gene-specific dispersion factor *𝛼_i_*, which relates the observed variance on the quantification and the average value for the particular gene *i*. The mean is the result of the quantity *q_ij_*, which is proportional to the expected read counts, and the size factor *s_j_*, identical for all the genes in a sample to account for variability in depth of sequencing. The design matrix *x_j_* indicates the condition on sample *j* and its covariates, and the GLM returns the 𝛽_𝑖_ coefficients. These latter coefficients are used to calculate the log2 fold-changes in gene expression between conditions. DESeq2 divides this process into three: i) estimate size factors on each sample *j*; ii) estimate dispersion factors 𝛼*_i_*; iii) adjust the negative binomial GLM 𝛽_𝑖_ coefficients and Wald statistics with a Wald test. DESeq2 applies a second filtering step via an “*independent filter*” under the “*result*” function to reduce false positive results by using the normalised counts to discard genes with no chance of being significant before even running the Wald test. This increases the detection power by decreasing the type I error rate, reducing the number of false positives and the number of tests, optimising the adjusted P-values. The P-values adjustment of the DGE results is derived by an adapted Benjamin-Hochberg (BH) method (P-values are multiplied with the number of tests and then divided by the rank, resulting in the adjusted p-value).

Principal Component Analysis (PCA) was performed using the raw count matrix after filtering, in which the feature space is given by the genes and the points are the samples. The covariates included in this analysis were ‘gender’, ‘age’ and ‘familyID’. The first and second PC were plotted to visualise if there were groups of samples that cluster together.

The MA plot is used to visualise the difference in gene expression between two conditions by

showing the log fold change (M) on the y-axis against the average of normalised counts (A) on the x-axis, where each dot represents a gene. The MA plot provides an overview of potential differentially expressed genes and enables a visual check on the distribution of data. Genes with a low average count usually present higher variability in fold change, which gives the graph a typical trumpet-shape pattern.

**1.5 DEGs in brain expressed tissues**

The Genotype-Tissue Expression (GTEx) database (v. 8) (https://gtexportal.org/home/) was used to check the correlation in gene expression between brain tissue and lymphoblastoid lines: the data-set can be found at ‘GTEx Analysis V8; RNAseq data; Median gene-level TPM by tissue’. Median expression was calculated from the file ‘GTEx_Analysis_2017-06-05_v8_RNASeQCv1.1.9_gene_tpm.gct.gz’. The tissue sources used in this analysis are all ‘brain tissues’, the ‘whole blood’ and the ‘EBV transformed lymphocytes’. The ‘average brain tissue’ expression was calculated from the expression across 13 different brain regions: amygdala, anterior cingulate cortex (BA24), caudate, cerebellar hemisphere, cerebellum, cortex, frontal cortex (BA9), hippocampus, hypothalamus, nucleus accumbens, putamen, spinal cord, and substantia nigra. The expression of the DEGs was assessed using the recommended TPM thresholds, considering basal gene expression when the TPM value is above 0.1.^2^

**1.6 DEGs validation**

The validation from the DGE analysis was performed through: i) An additional RNAseq dataset from the PsychEncode Consortium BrainGVEX RNAseq Study;^3^ ii) Gene-based association analysis via MAGMA version 1.9,^4^ using GWAS summary statistics of BD (41,917 BD; 371,549 controls);^5^ iii) Polygenic priority score (PoPS),^6^ which prioritizes candidate genes combining genetic associations and functional annotations.

The PsychEncode dataset (Project SynID: syn4921369) comprised 73 BD patients and 259 controls derived from human brain tissue (BA9). Five control samples and two BD patients were excluded based on ancestry criteria, and two additional control samples were excluded during QC processing. The analysis followed the same pipeline used for the main analysis (eFigure 1), but as PsychEncode is a case-control rather than a family-based cohort design, only ‘gender’ and ‘age’ were set as covariates.

Gene-based association analysis was performed via MAGMA version 1.9,^4^ using GWAS summary statistics of BD (41,917 BD; 371,549 controls).^5^ This analysis excluded *PPP2R3B*, which maps in the pseudoautosomal region of sex chromosomes, for which SNPs were not available in the GWAS dataset.

Fifteen out of the 59 remaining DEGs were nominally significant according to MAGMA gene-based analysis: *RC3H1, MRPS27, OTUD6B, XPO5, MYO18A, GPT2, PFKFB2, NRM, GNB2, PPDPF, DNAJA2, RIPOR1, CEBPZ, ENSG00000229127* and *ENSG00000279277* (eTable 5). The PoPS score is based on MAGMA gene-based P-values and a matrix of biological features including curated pathways, expression patterns, and protein interaction data. The provided feature matrix lacks non-protein coding genes, and for this reason nine genes were excluded for this analysis: *ENSG00000279277*, *ENSG00000229127*, *ENSG00000256482*, *OVCH1-AS1*, *ENSG00000227757*, *ENSG00000224610*, *ITGB2-AS1*, *LINC01829* and *LINC01237*. PoPS scores higher than 0.2 are interpreted as evidence of causality. The lncRNA *ENSG00000279277* resulted the most significant gene from the MAGMA analysis (gene-based P-value= 1.58E-07), and the only gene-based significant hit amongst those with no PoPS score.

**1.7 WGCNA pipeline**

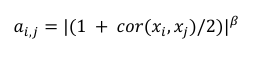
WGCNA uses graph theory to construct a network where genes are nodes, and the strength of co-expression between them is represented by edges. The method groups highly interconnected genes into modules, which are sets of genes with similar expression patterns. The module’s eigengene can be used in downstream analyses, such as correlation with external traits or identification of module membership of individual genes. As WGCNA is very sensitive to noise, a stringent filter was applied to raw counts matrix to include only those gene features with at least 10 count reads in at least 90% of our sample (28 individuals). A signed network is built by clustering gene outcomes with a positive correlation in gene expression patterns. Conversely, an unsigned network would make clusters of gene expression patterns with absolute values of correlations.^7^ An adjacency matrix is built by exposing the signed similarity matrix to a soft threshold (𝛽) power as follows:

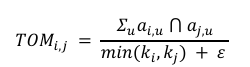
The signed similarity matrix measures the correlation between the expression profile of gene *i* and gene *j*, covering a range of values from 0 to 1. The soft threshold increases strong correlations and reduces those that are weak. This parameter was chosen based on the scale-free topology criterion and set to 12. The adjacency matrix is then transformed into a Topological Overlap Matrix (TOM) as follows:

TOM accounts for indirect connections and shared neighbors, which considers the overall connectivity patterns rather than only taking into account direct correlations, improving preciseness and robustness.^8^ The *k_i_* represents the total connectivity, which sums all the connections of the gene *i* to the network. The numerator measures the number of shared neighbors between both genes and the denominator represents the minimum number of neighbors between them, plus 𝜀 (pseudocount) to avoid division by 0. *TOM_ij_* ranges from 0 to 1, where higher values indicate stronger topological overlap. The complementary of the TOM matrix, or dissimilarity matrix (1-TOM), is used in a clustering algorithm such as Dynamic Tree Cut to detect the modules present in the network. The Dynamic Tree Cut constructs a hierarchical tree and cuts it at a certain height: it makes progressively higher cuts and checks the module eigengene-based connectivity as a reference of quality in an iterative process to automatically identify the optimal number of modules. The network was manually constructed in two steps, by setting an initial cut with a minimum module size of 15 genes and then merging modules with 75% correlation, according to WGCNA manual recommendations. The module eigengene (*E*), which is the first principal component of a module, represents its gene expression profile. It can be correlated to the condition status (affected/unaffected) to assess co-expressed genes related to the affection status. The module membership (MM), or eigengene-based connectivity *K*, measures the correlation between the expression profile x of gene *i* and eigengene *E* of module *q* as follows:

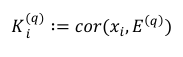

The most correlated module with the trait was inspected for networks of physical interactions amongst the module genes using the Ingenuity Pathway Analysis (IPA, [www.ingenuity.com](http://www.ingenuity.com)). IPA computes a score based on the probability of finding a given pool of genes in a network from the Ingenuity’s Knowledge Database by chance (score¼ log [Fisher exact test probability]).

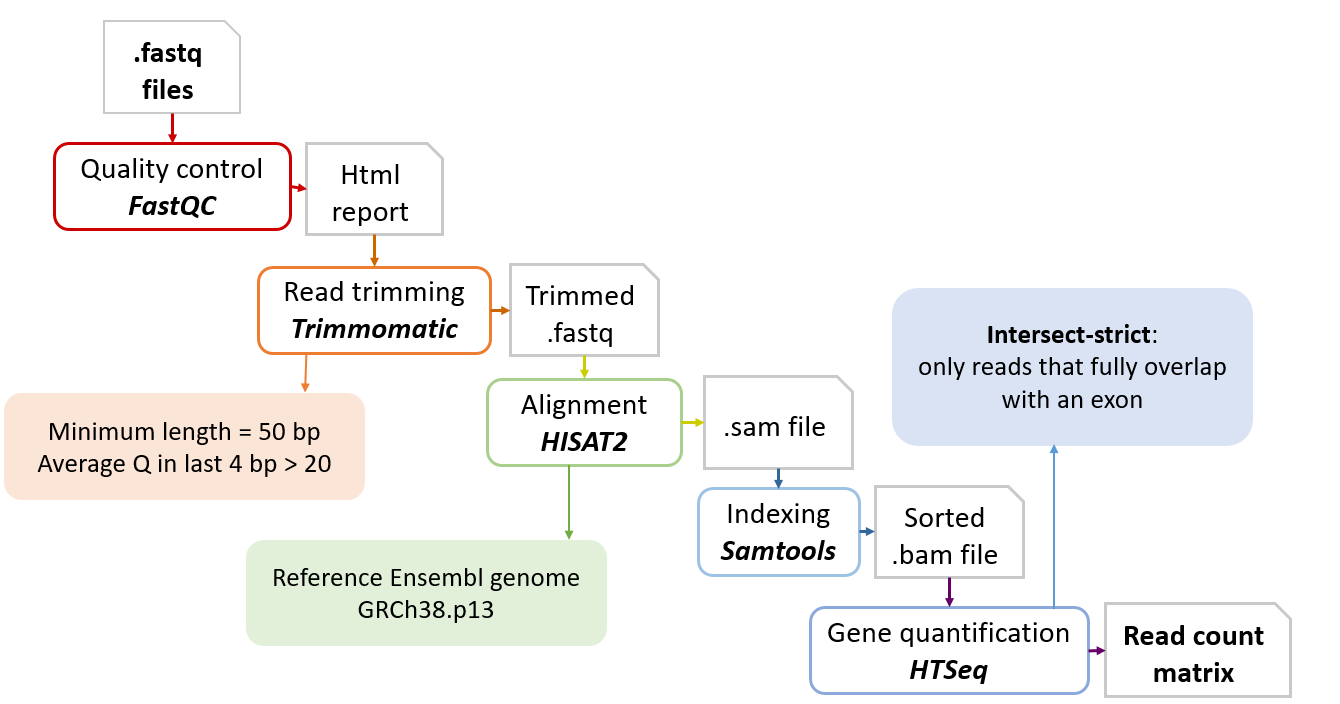

**eFigure 1:** The workflow of the RNAseq processing pipeline is shown. The *.fastq* files were checked for quality, trimmed, and aligned against Ensembl reference genome GRCh38.p13. The indexed reads are counted to create a raw count matrix using HTseq in *intersect-strict* mode.

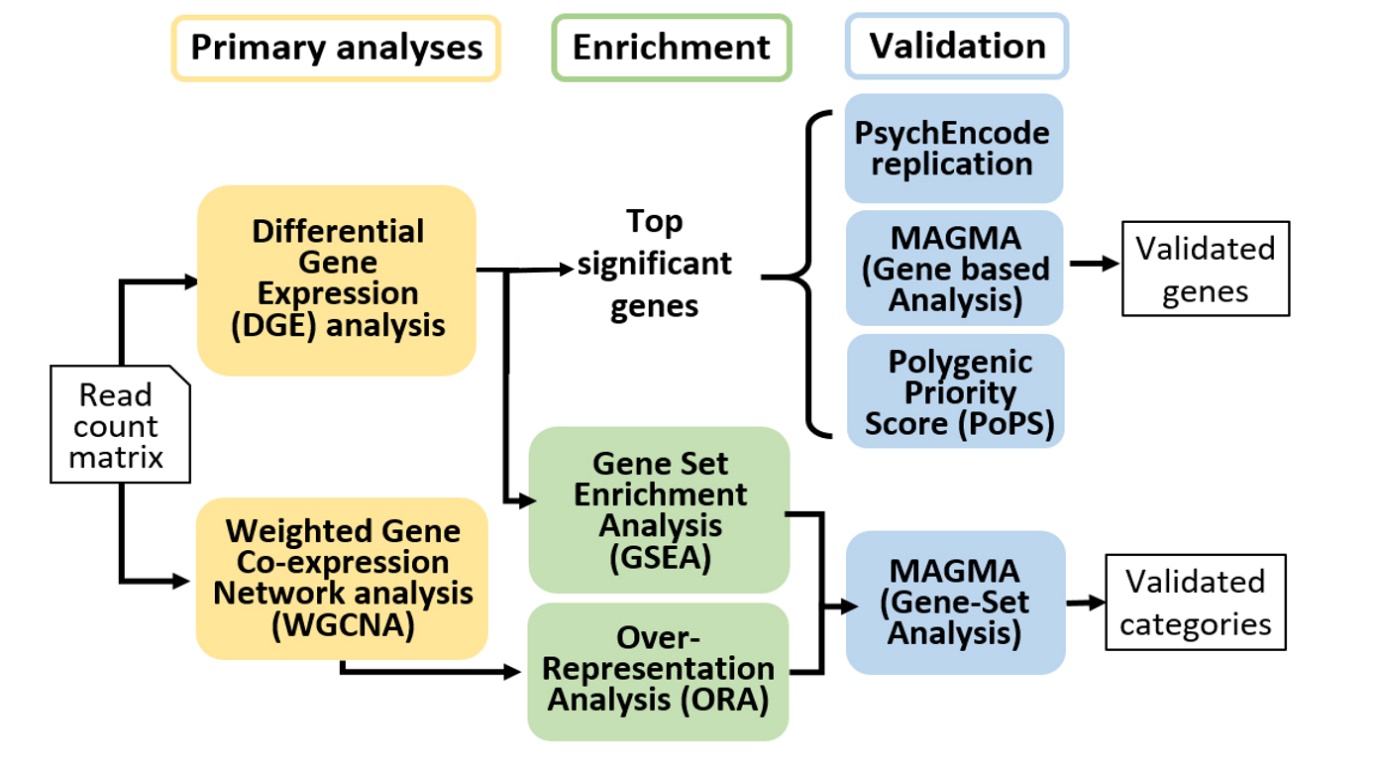

**eFigure 2:** Complete workflow of the present study. Two primary analyses were performed from RNAseq read counts: i) Differential Gene Expression (DGE) analysis; and ii) Weighted Gene Co-expression Network Analysis (WGCNA). The second step consists of enrichment analyses as follows: gene set enrichment analyses (GSEA) using as input file the DGE results of 15,231 genes and over-representation analysis (ORA) for individual correlated modules from the WGCNA. The validation of our results was performed: i) For individual DEGs using a replication study of 73 BD patients and 259 controls derived from human brain tissue (BA9) from the PsychEncode Consortium, a MAGMA gene-based analysis of the latest GWAS summary statistics in BD (41,917 BD; 371,549 controls), and a polygenic priority score (PoPS); ii) The enriched categories from GSEA analysis or significant modules of the WGCNA were validated through the MAGMA gene-set analysis.

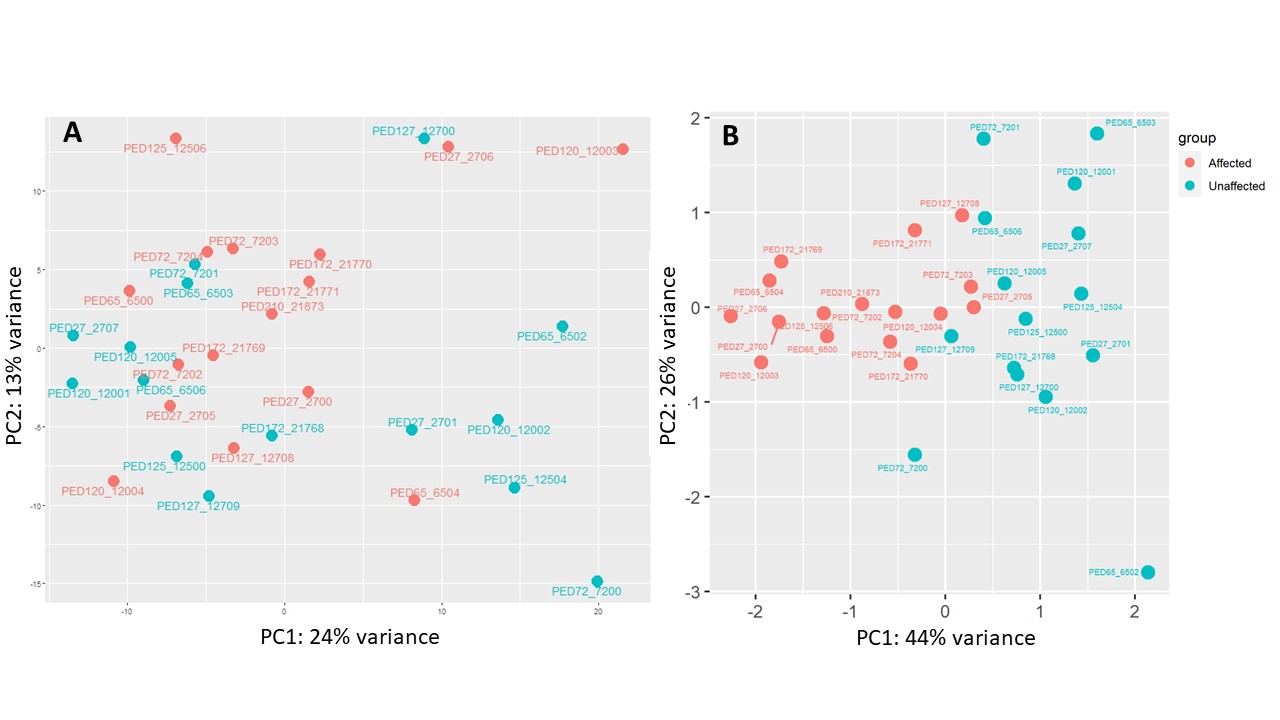

**eFigure 3:** PCA plots of count matrix of genetic features in the 31 samples, comprising n=16 BD patients and n=15 unaffected relatives. The PCA analysis accounted for ‘gender’, ‘age’, and ‘family’ effects. A) The PCA performed with the total number of 28,525 genetic features, where the first component explains 24% of the variability, while the second component explains 13% of the variability; B) The PCA performed only with the 60 DEGs identified from this study, showing that affected and unaffected relatives are clustered in two distinct groups.

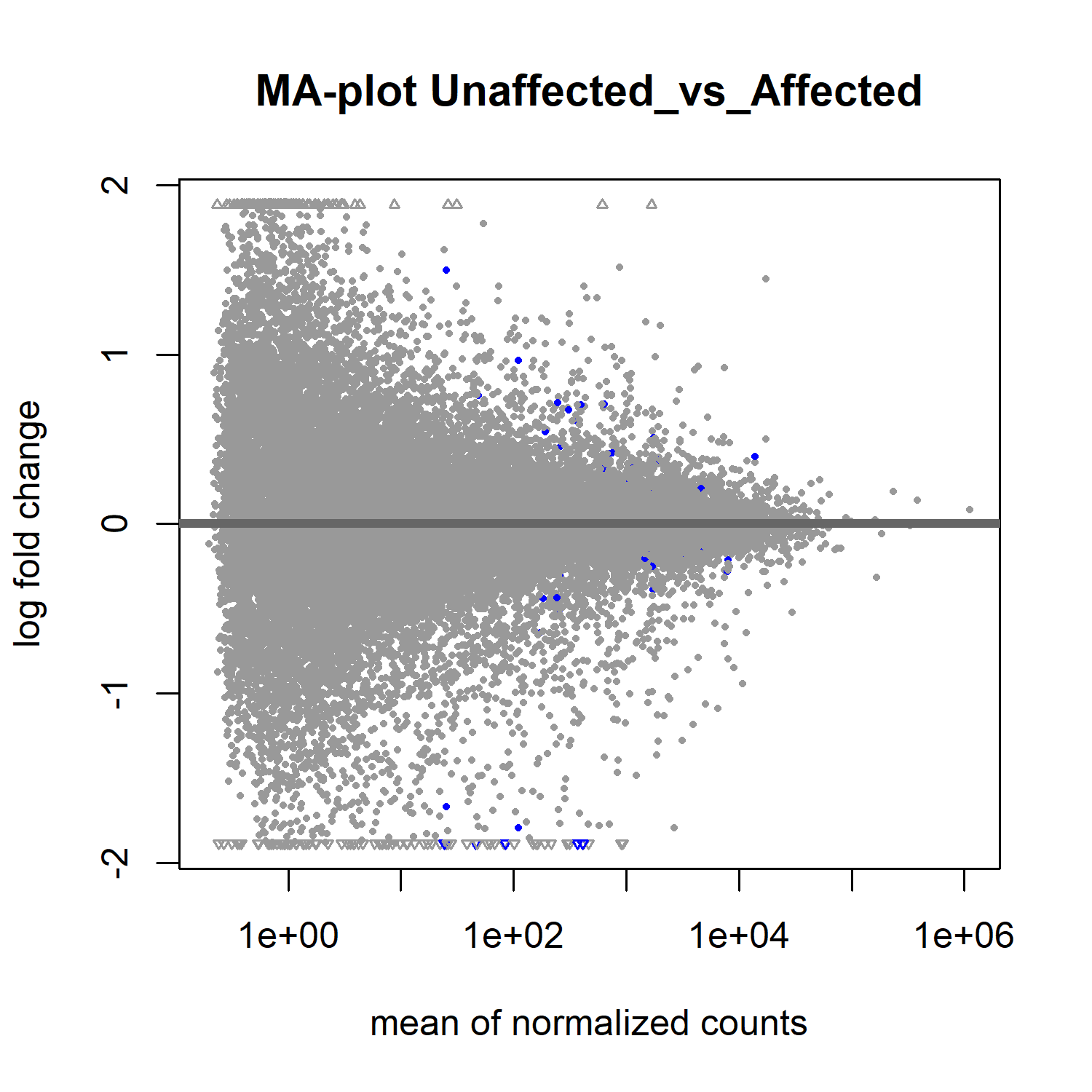

**eFigure 4:** MA plot for differential expression analysis. Mean of normalised counts per gene (A values) are represented in the x-axis, and log fold-change (M values) are represented in the y-axis, where each dot represents a gene. Significant gene expression changes (adjusted P-value <0.05) are depicted in blue.

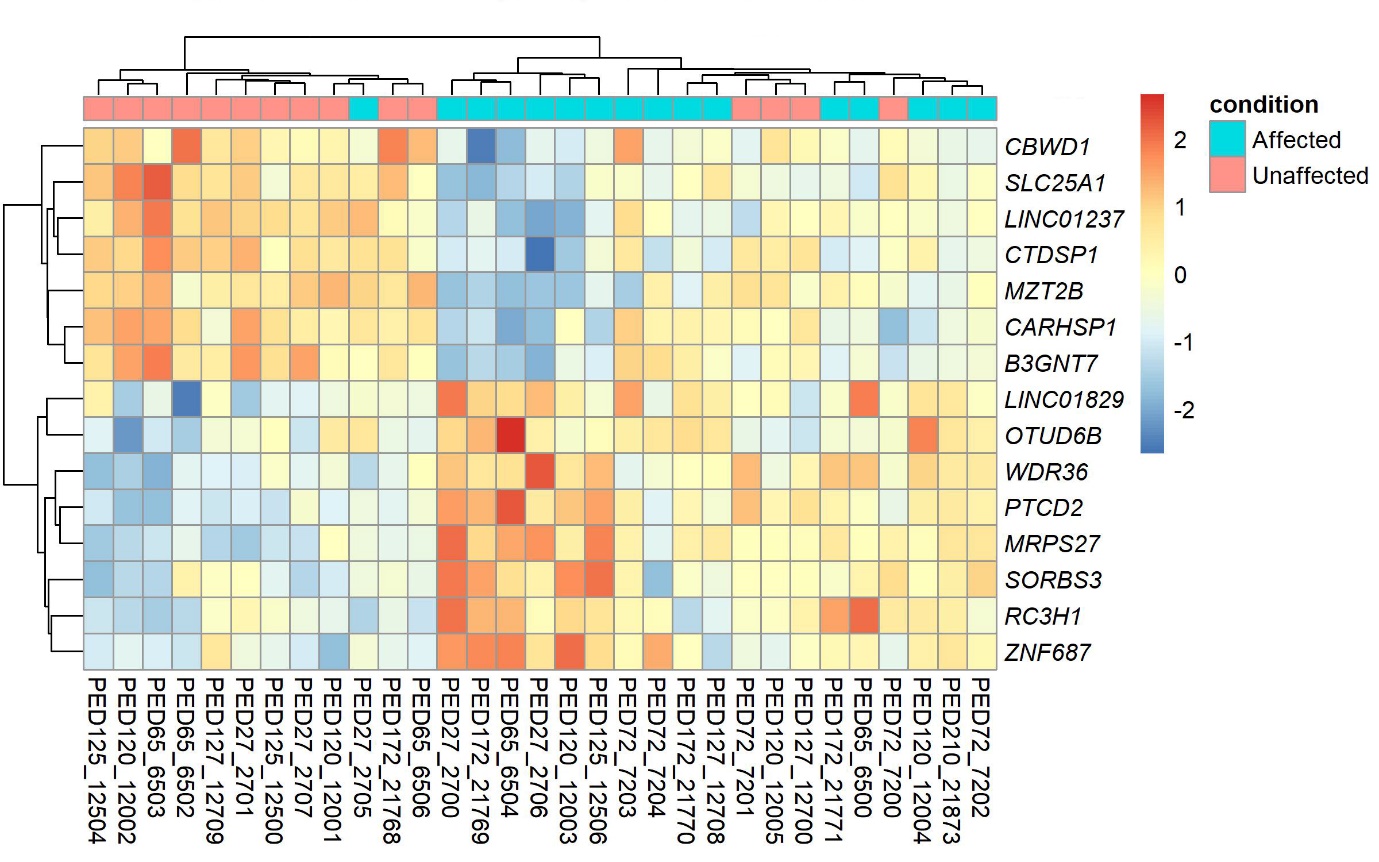

**eFigure 5:** Heatmap of DEGs with adjusted P-value <0.01 using the 31 individuals of this study. Hierarchical clustering using expression of these 15 genes is able to parse the individuals into two groups, principally corresponding to affected (depicted in turquoise) and unaffected relatives (depicted in pink). The gene expression, in log2 fold change of normalised counts, ranges from red (higher) and blue (lower).

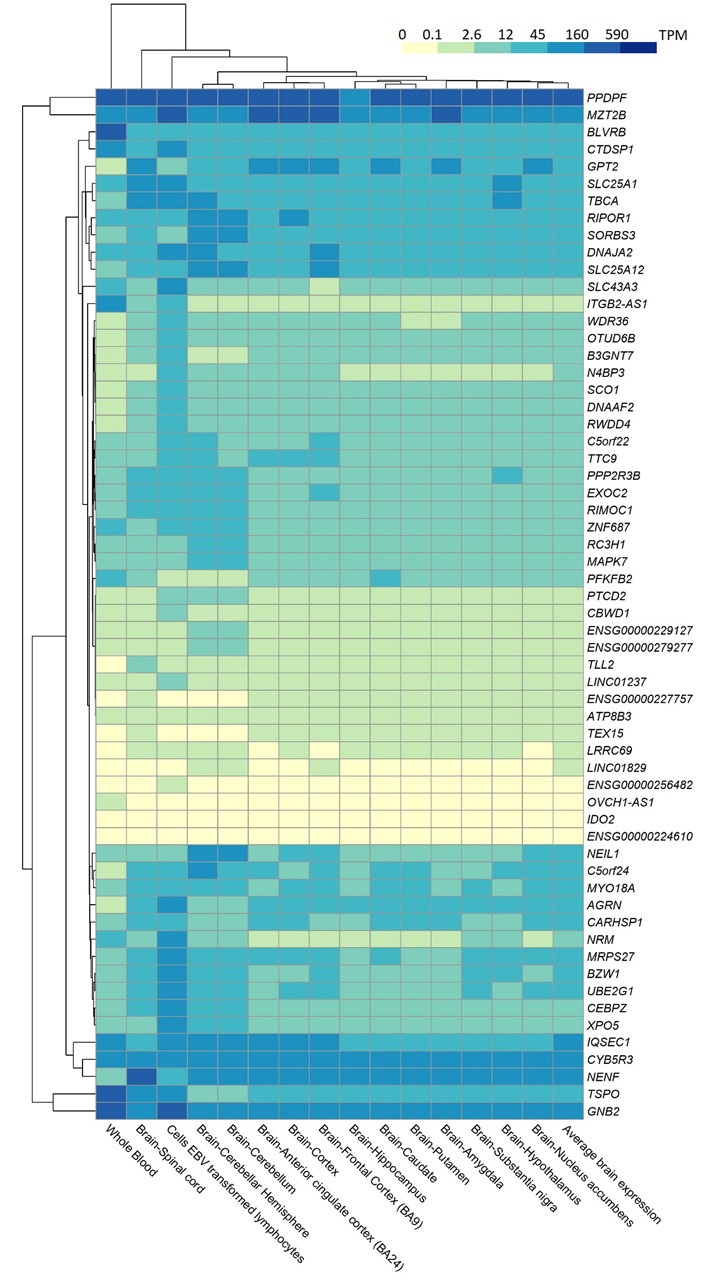

**eFigure 6:** Gene expression of the 60 significant DEGs across whole blood, lymphoblastoid cell line, 13 brain tissues, and their TPM average values using the GTEx v.8 dataset. TPM values lower than 0.1 are in yellow, showing no basal gene expression, while higher expression is depicted in blue shades. Genes and tissues are grouped using hierarchical clustering.

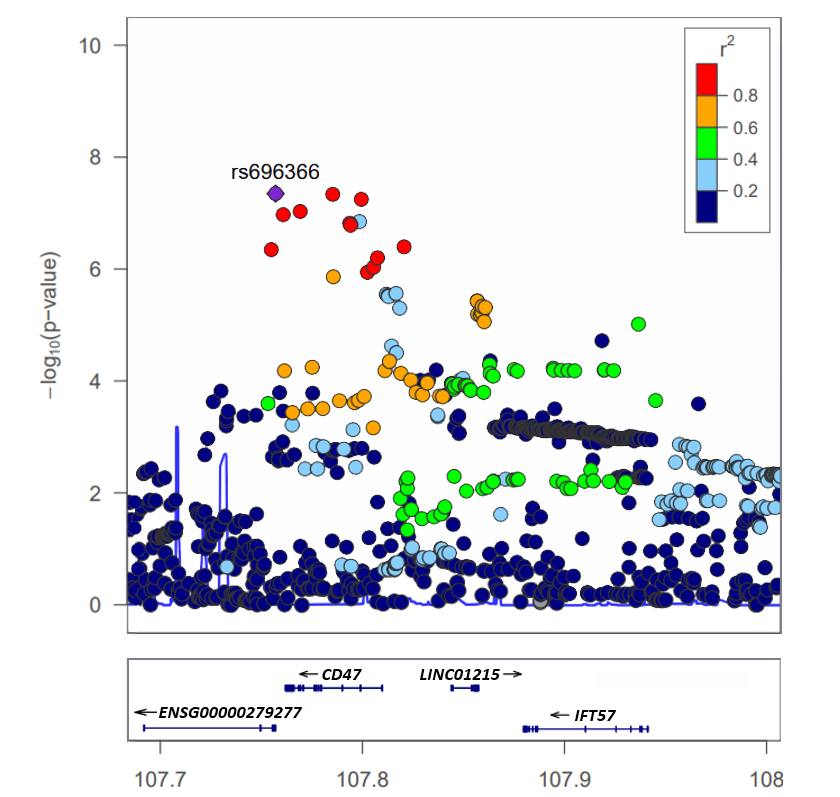

**eFigure 7:** Association plot of *ENSG00000279277* in the GWAS summary statistics of Bipolar Disorder of the PGC, according to the GRCh37/hg19. The y-axis indicates the significance of association as the negative logarithm of the P-value (–log P-Value), and x-axis indicates the physical position along the gene in megabases (Mb). The name of the most significant SNP, rs696366 which resides in intron 1 of *ENSG00000279277,* is indicated. *Linkage disequilibrium* (r^2^) between SNPs is calculated and plotted in different colours, whose ranges are in the above part of the plot.

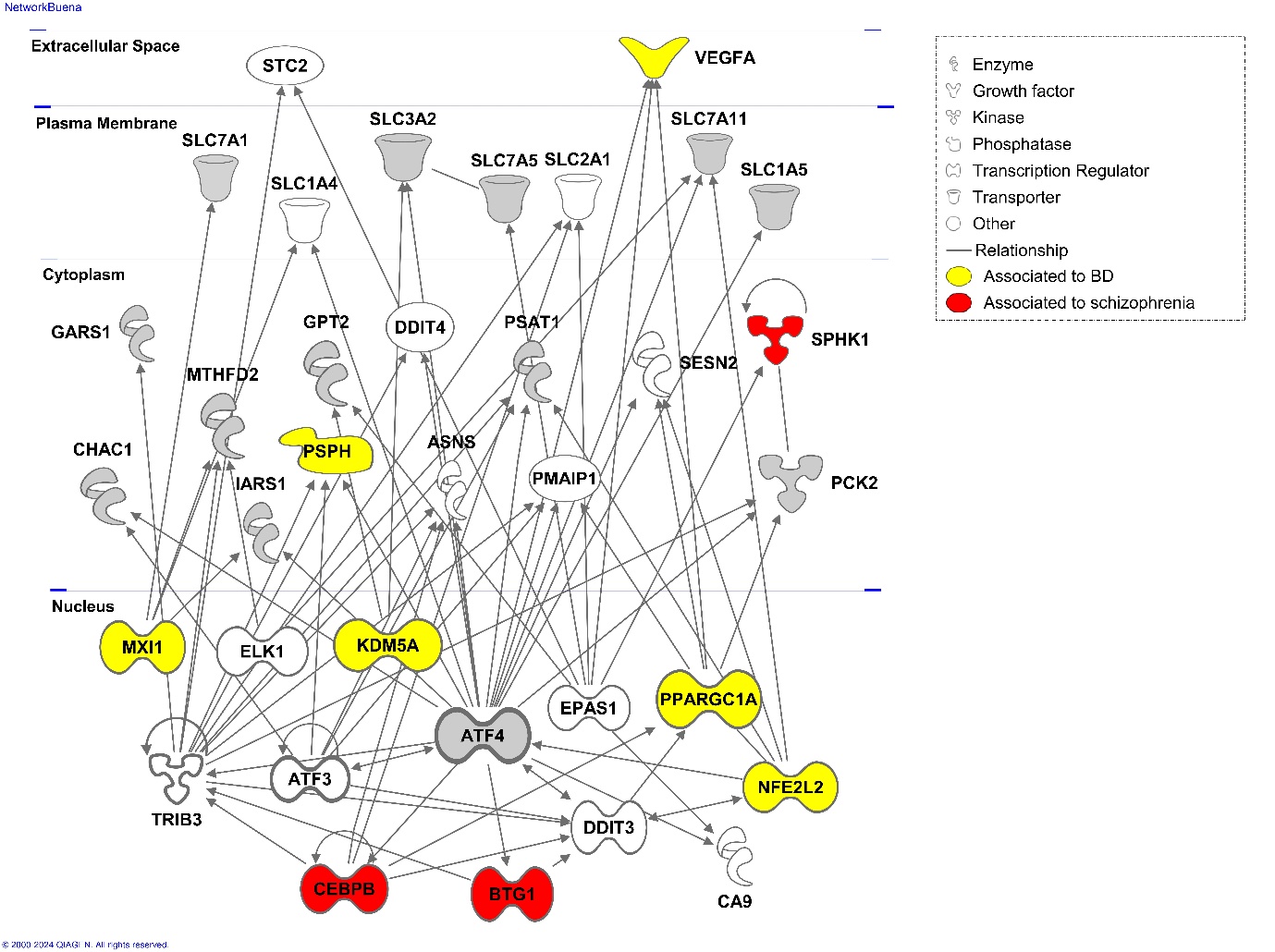

**eFigure 8:** Protein-protein interaction network via IPA was performed using the list of genes (N=98 genes) from the *darkolivegreen4* module, which showed the most significant enriched categories after validation. The figure shows the top network (scored 32), amongst the three networks identified, which is related to amino acid metabolism and small molecule biochemistry. Proteins in yellow and red represent genes associated with BD or schizophrenia, respectively, according to PGC GWAS studies.^5^ The P-values from gene-based analysis using PGC GWAS datasets are as follows: *VEGFA* (*P*=0.046), *SPHK1* (*P*=0.00011), *PSPH* (*P*=0.039), *MXI1* (*P*=0.0099), *KDM5A* (*P*=0.019), *PPARGC1A* (*P*=0.000078), *NFE2L2* (*P*=0.014), *CEBPB* (*P*=0.021), *BTG1* (*P*=0.026). Proteins in grey represent genes from the *darkolivegreen4* module. Up-regulatory effects are represented by outward pointing arrows, down-regulatory effects are represented by outward ticks, and circular arrows indicate homotypic interactions.

**Supplementary Methods References**

1.	American Psychiatric Association. Diagnostic and Statistical Manual of Mental Disorders, Fourth Edition. Washington, DC. 1994.

7.	Zhang B, Horvath S. A general framework for weighted gene co-expression network analysis. Stat Appl Genet Mol Biol. 2005;4:Article17.

8.	Khatri P, Sirota M, Butte AJ. Ten years of pathway analysis: current approaches and outstanding challenges. PLoS Comput Biol. 2012;8(2):e1002375.
